# Progressive changes in descriptive discourse in First Episode of Schizophrenia: A longitudinal computational semantics study

**DOI:** 10.1101/2021.09.29.21264300

**Authors:** Maria Francisca Alonso-Sánchez, Sabrina D. Ford, Michael MacKinley, Angélica Silva, Roberto Limongi, Lena Palaniyappan

## Abstract

Computational semantics, a branch of computational linguistics, involves automated meaning analysis that relies on how words occur together in natural language. This offers a promising tool to study schizophrenia. At present, we do not know if these word level choices in speech are sensitive to illness stage (i.e. acute untreated vs. stable established state), track cognitive deficits in major domains (e.g. cognitive control, processing speed) and relate to established dimensions of formal thought disorder. Here we study samples of descriptive discourse in patients with untreated first episode of schizophrenia (x□ 2.8 days of lifetime daily dose exposure) and healthy subjects (246 samples of 1-minute speech; n=82, FES=46, HC=36) using a co-occurrence based vector embedding of words. We obtained six-month follow-up data in a subsample (99 speech samples, n=33, FES=20, HC=13). At baseline, the evidence for higher semantic similarity during descriptive discourse in FES was substantial, compared to null difference (Bayes Factor =6 for full description; 32 for 10-words window). Moreover, the was a linear increase in semantic similarity with time in FES compared to HC (Bayes Factor= 6). Higher semantic similarity related to lower Stroop performance (accuracy and interference, response time), and was present irrespective of the severity of clinically ascertained thought disorder. Automated analysis of non-intrusive 1-minute speech samples provides a window on cognitive control deficits, role functioning and tracks latent progression in schizophrenia.

## 1. Introduction

Language disorganisation is a prominent feature in psychosis, and it is commonly encountered as a disorder in generating interpersonal discourse. This produces a significant functional impairment especially as it interferes with one’s ability to describe or explain attributes and thus socialise in everyday life ^1^. When having a descriptive discourse that describes a concrete referent such as a picture to a second person, patients with schizophrenia make unusual word choices ^2^, exhibit repetitiveness and convey less information (referred to as ‘weakening of goal’^3^ or ‘poverty of content’^4^) than healthy controls^3,5^. In particular, the restricted repertoire of word selection, characterised by smaller loops of word-to-word connectivity that occurs with more proximal repeats in selected words, becomes apparent even before overt psychosis^6^, predicts later onset of psychosis^6,7^, and becomes more pronounced during the first episode^7^, and relates to reduces social and occupational functioning^8^.

Descriptive discourse involves multiple levels of cognitive processing ^9^ to integrate parts and attributes to the whole to produce a descriptive schema^10^. We often employ descriptions in the service of rhetorical functions (i.e., ways to inform, argue, persuade someone) through our choice of words. In psycholinguistic terms, descriptive discourse requires semantic competence^1^ and appropriate lexical access to a connectionist system of word organised by their conceptual relation with one another ^10^. In this context, lexical units (words) with a higher likelihood of occurring together have a stronger connection or a smaller distance between them (distributional semantics) ^11^. This idea follows the original spreading-activation hypothesis of lexical representations in the brain ^12^. Competitive theories of lexical selection assume that lexical representations must overcome interference from the neighbour’s activation through lateral inhibition ^13^. Applying this to the picture description task, a failure of appropriate selection via inhibition at lexical level may give rise to a description that is replete with words that are highly associated with each other, without capturing the different attributes of the picture at hand.

A proactive ‘top-down’ contextual guidance during discourse can reduce the overreliance on the bottom-up activation of the lexico-semantic network for word selection ^14^. A breakdown in this contextual guidance, implemented as top-down inhibition from inferior frontal to semantic storage systems ^15^, has been variously described in schizophrenia ^16^. A large body of literature demonstrates frontal cognitive control deficits in schizophrenia, exemplified by reduced performance in color-word Stroop Task that tests one’s ability to inhibit competing semantic categorical representations when making a choice ^17^. In particular, the increased Stroop interference effect, in both response time and accuracy measures, has been interpreted as a marker of impaired inhibitory aspect of cognitive control ^17^. Abnormalities in this aspect of cognitive control has been previously related to conceptual disorganization, a symptom related to linguistic aberrations in schizophrenia ^18,19^. On this basis, we can expect cognitive control deficit to influence the word selection during a descriptive discourse in schizophrenia.

When examining similarity among the words used during a discourse, there are broadly 2 approaches. One approach is to count the instances of the repetition of a word. This phenomenon is described as perseveration in clinical rating scales ^3,4^. A measure of lexical diversity called Type-Token Ratio (TTR; ratio of unique to total words in a text) is computed based on such repetitions. As exact repetitions are relatively rare, perseveration is often not detectable in cross-sectional interviews ^20,21^, and results from TTR studies are inconclusive in schizophrenia ^22–24^. Graph theoretical approaches that rely on the proximity between two repetitions, rather than counting the instances of repetitions, appear to carry diagnostic and prognostic information in schizophrenia ^8,25^. But this approach cannot distinguish meaningful repetitions of informational value (e.g., “He liked the idea of travel, and the memory of travel, but not travel itself”) from the problematic repetitions that affect communication. The second approach is to employ distributional semantics to estimate the similarity, rather than exact repetition, among a set of words. This taps on a network based distributional model of words. If lexical units are interconnected based on their co-occurrence in everyday language, then similarity among a set of words used during a discourse can be quantified on the basis of this distributional co-occurrence.

Approaches from distributional semantics have been applied to study the relationship among words produced during various speech elicitation tasks in schizophrenia. The most popular approach, introduced by Elvevåg ^26^, involves the use of latent semantic analysis (LSA) that taps on the document-level statistical co-occurrence of words in a large corpus of written texts; this determines their position in the semantic space based on the “company they keep”. The cosine similarity of this spatial index can then be computed among the words spoken by a patient. Several studies have demonstrated the potential utility of distributional semantics in predicting the onset of psychosis ^2,27,28^, examining thought disorder ^29–31^ and its neuroanatomical basis of linguistic disruptions in psychosis ^32^. Other similar methods improvised on LSA, by weighting the statistics of co-occurrence on the basis of the actual proximity of words in the sentences occurring in the reference corpora ^33–39^. We employ one such improvised method (CoVec), that has been employed previously in the study of semantic fluency tasks in schizophrenia^40,41^. Cosine similarity can be computed between words that are adjacent to each other within a window, indicating if words proximal to each other are sampled from a narrow semantic space. As spoken text rarely assumes the form of sentences, a finite moving window (e.g. 5, 10 or 20 words size^40–43^) is often used to measure this proximal similarity. Cosine similarity among the full frame of words in a descriptive text indicates the sematic diversity of all words employed to provide the complete description of a referent (e.g., a picture).

Studies employing distributional semantics have often employed the term coherence to describe the degree of similarity (e.g. local coherence^42^, semantic coherence^26^, or cohesion^44^) or incoherence when describing its pathological reduction^29,39^ (see ^33,45^ for a review). While a number of NLP studies have employed the term coherence in this sense, we use the term ‘similarity’ rather than coherence when employing cosine similarity here. Hoffman pointed out that coherence is a psychological experience of a listener and not a property of a text ^46^. To experience a text as coherent, the listener must employ a subjective interpretive synthesis that depends on their experience of the referent (i.e., drawing the linkage between the described object and the presented text) and directionality (i.e. which word or idea came first), in addition to the dependency among the lexical/semantic units. Furthermore, words with low probability of co-occurrence can be coherently juxtaposed in certain contexts, that may not be apparent from the text itself. Also, metadiscursive (frameshifting^46^) elements can improve coherence for a listener (e.g., changing topics by saying “to go on a tangent for a bit”). For these reasons, we do not infer semantic *coherence* but only *similarity* from the indices of distributional semantics employed here.

We hypothesize that when faced with the task of providing a description of an unfamiliar concrete referent^47^ (a picture), patients with schizophrenia will employ words with higher probability of semantic co-occurrence (similarity). This anomaly will be evident even in the untreated, first episode phase of illness and relate to failed cognitive control in patients. We anticipate that unlike healthy controls who will show a static level of similarity in their word choice over time, a progressive worsening over time will be seen among patients, in relation to their social functioning. Of note, we assembled a sample of acutely unwell, first episode patients with < 14 days of lifetime exposure to antipsychotics at baseline. These patients were then treated in an early intervention clinic and followed up at 6 months period to examine their discourse stability. This allowed us to relate treatment variables (antipsychotic exposure) as well as outcome variables (SOFAS scores) to word similarity measures over time.

## 2. Results

### Demographic and clinical characteristics

Healthy controls and FES (First episode of schizophrenia) did not significantly differ in age, gender distribution or educational level. In the FES group, 20% of the participants were first generation immigrants (determined from self-report) while in the matched HC group this was 30%. There was no group difference in the use of English as first language (82% FES and 88% HC had English as first language). All the participants had English as their only transactional language. As expected, HC group performed better on a modified digit-symbol substitution task (DSST) measuring processing speed and Color-Word Stroop task. Clinical and demographic characteristics are provided in Table 1.

**Table 1.**
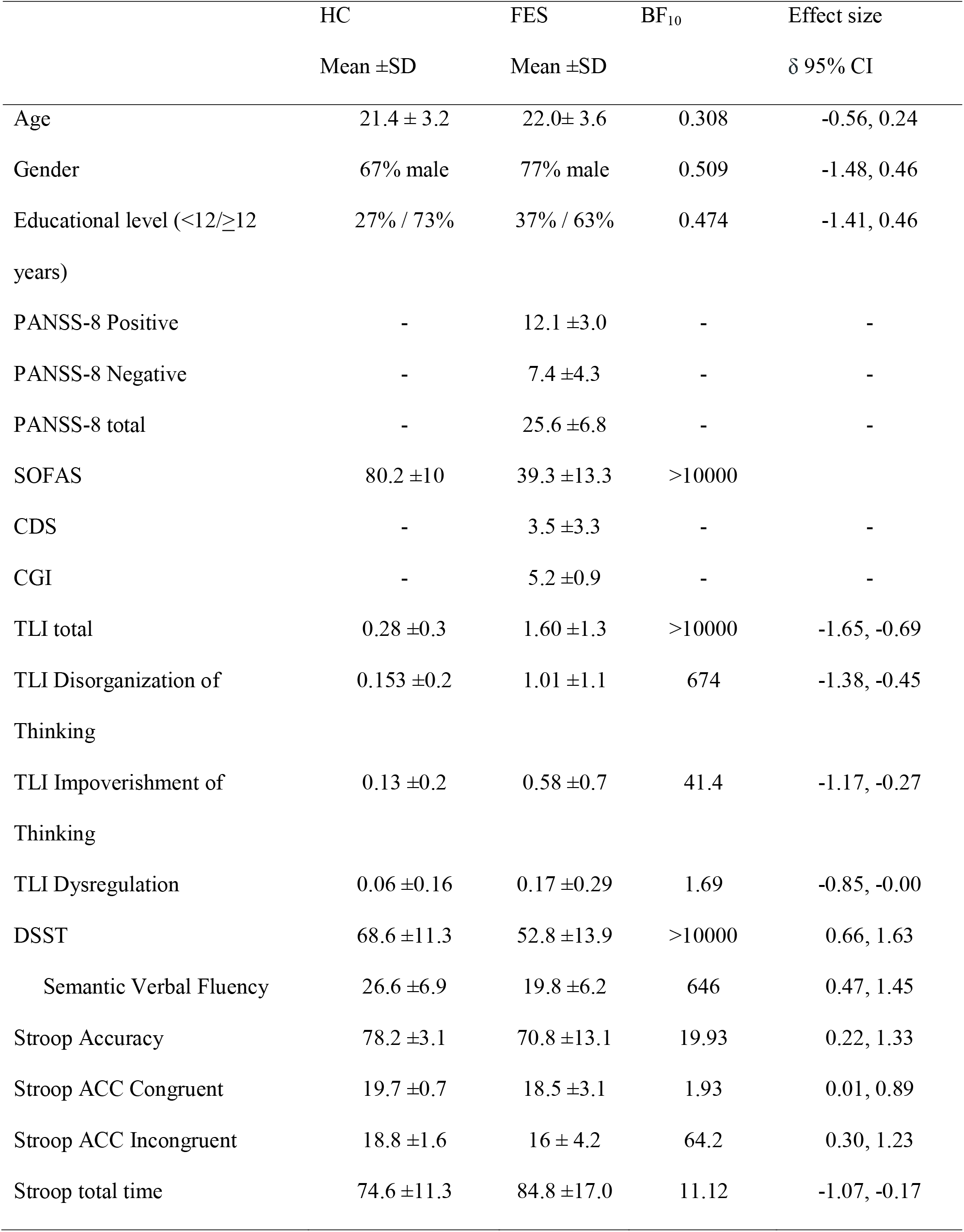
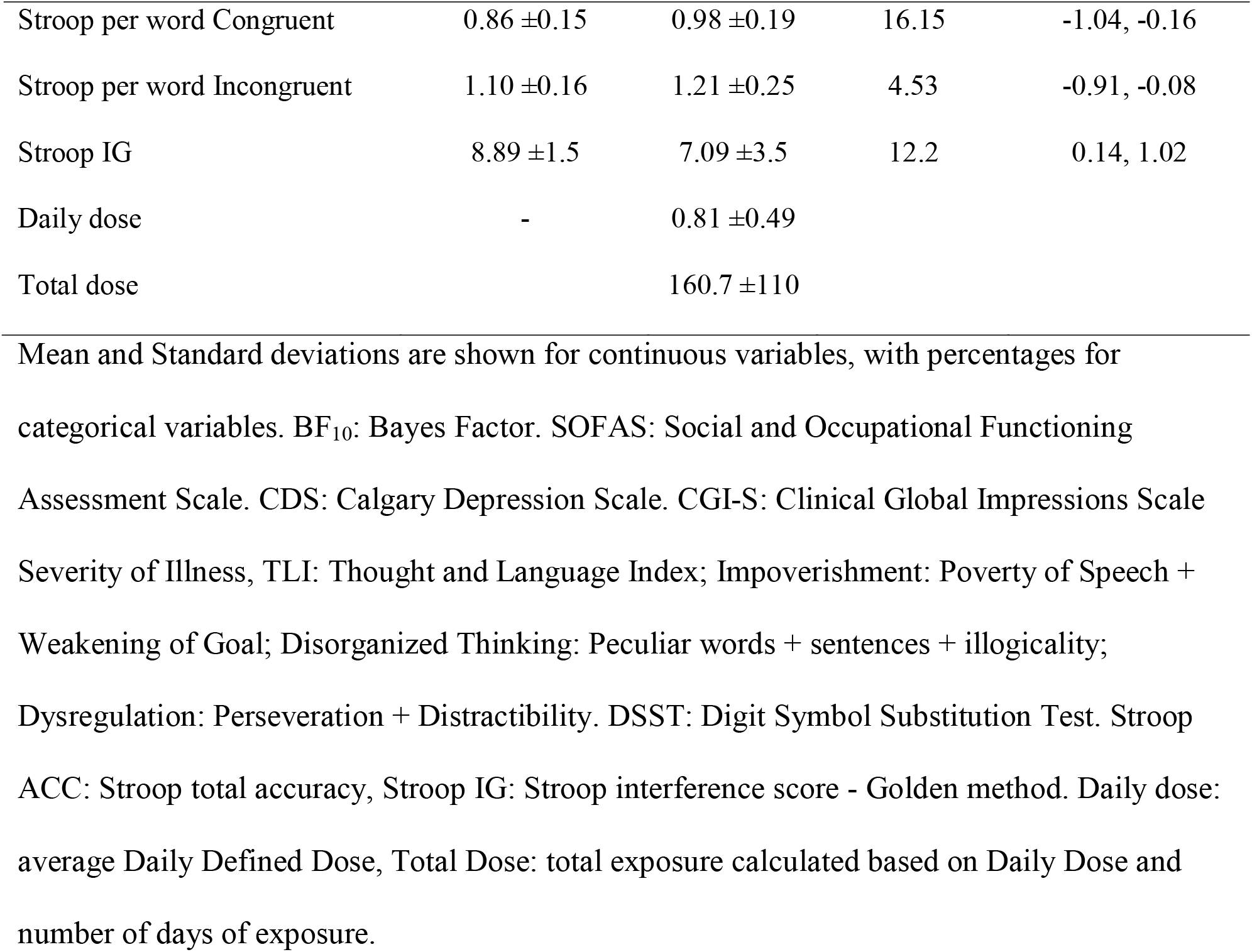

### Baseline differences in word similarity

In the description task, the groups did not differ in the number of words, but had higher similarity in the full frame (ASW-F; moderate degree of evidence against null) as well as in the 10-words frame (ASW-10; very strong evidence against null) compared to the HC group. These results are shown in Table 2 and Figure 1. To test if this increased word similarity was specific to the picture description task where word choices depend on the discursive discourse, we studied similarity of word choices in a category fluency task (participants were ask to produce as many animals as possible in one minute) from a subsample of subjects (HC n = 33, FES n = 39). There was no difference among groups in the mean similarity of generated words (HC: 0.497± 0.04; FES: 0.477± 0.05, BF_10_ = 0.696), indicating discourse-related specificity of increased semantic similarity in schizophrenia.

**Table 2.** Summary group differences at baseline

|  | HC | FES | BF<sub>10</sub> | Effect size |
| --- | --- | --- | --- | --- |
| | Mean ±SD | Mean ±SD | | $\delta$ 95% CI |
| NW | 70.6 ±14.9 | 68.4 ±30.3 | 0.249 | -0.32, 0.48 |
| ASW-F | 0.334 ±0.025 | 0.352 ±0.034 | 6.53 | -1.05, -0.17 |
| ASW-10 | 0.400 ±0.023 | 0.421 ±0.031 | 32.76 | -1.14, -0.25 |

NW: Number of words, ASW-F: Average Similarity of Words full frame, ASW-10: Average Similarity of 10 words frame. Note that the variables reported here are individually averaged across 3 speech samples per subject. BF<sub>10</sub>: Bayes Factor.

**Figure 1:**
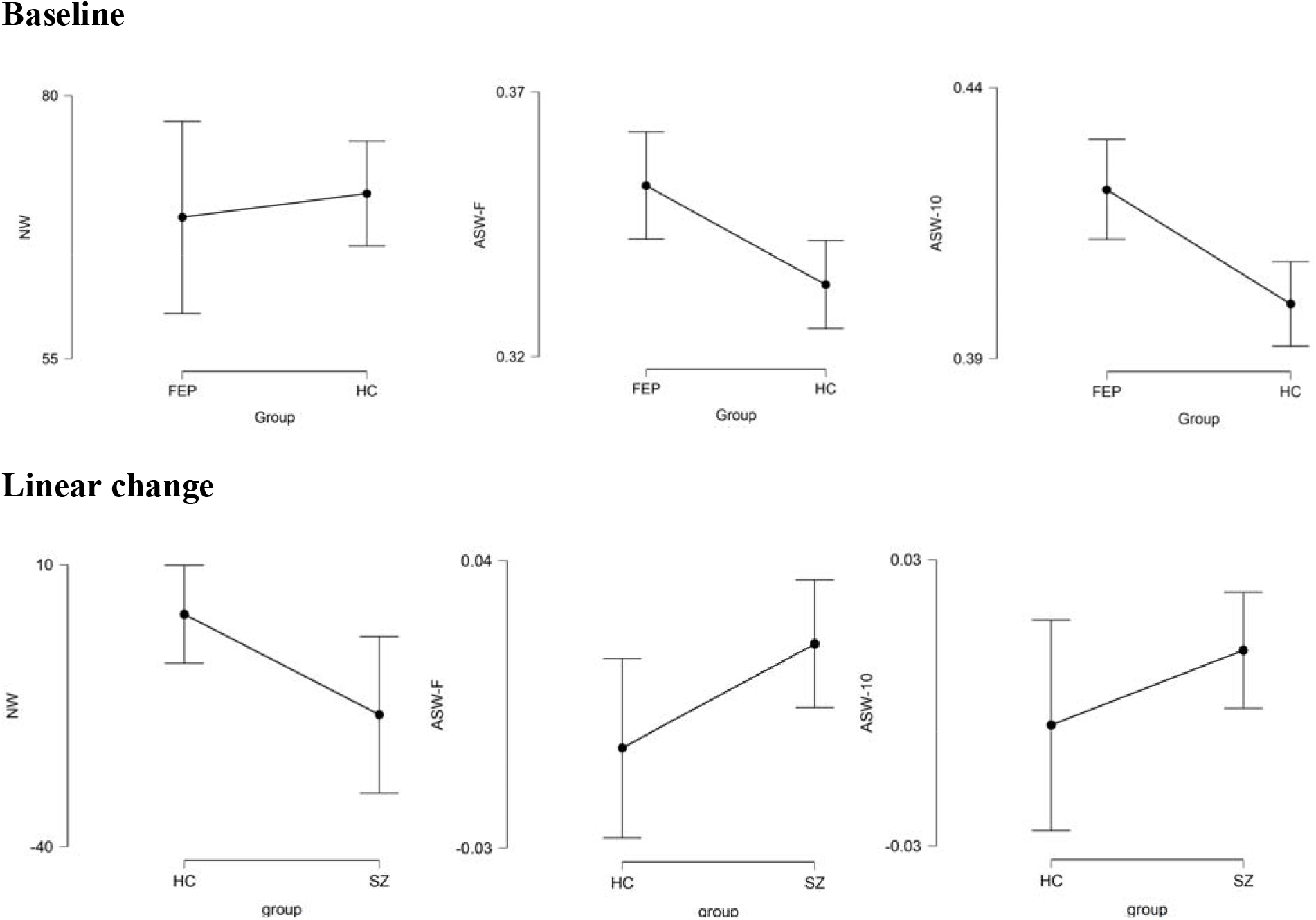
Group differences in linguistic variables at baseline and the change over time of linguistic variables Descriptive plots of 95% credible interval between groups. NW: Number of words. ASW-F: Average Similarity of Words full frame. ASW-10: Average Similarity of Words 10 words moving window. FES: First Episode of Schizophrenia. HC: Healthy control. SZ: Group with Schizophrenia.

### Longitudinal changes in word similarity

In the 6-months follow-up sample (n=33, FES=20, HC=13), the 2 groups were matched for age (SZ: 22.5± 5.0; HC: 21.5± 3.1, BF_10_ = 0.390) and gender (SZ: 80% male; HC: 70% male, BF_10_ = 0.611). Patients with SZ had strong evidence for functional improvement based on SOFAS scores (Baseline: 41.5± 13.5; follow-up: 61.0± 12.9; mean change = 19.5 ±14.3; BF_10_ for paired t test = 4868), and clinical improvement based on a reduction in PANSS-8 total score (Baseline: 25.2± 5.7; Follow-up: 15.1± 5.0, mean change = -10.25 ±4.9; BF_10_ for paired t test >10000) from baseline to follow-up assessment, as expected following clinical intervention (medication doses detailed below). While positive symptoms score improved (Baseline: 12.5± 2.6; Follow-up: 5.2± 1.7, BF_10_: >10000), the negative symptoms of the PANSS did not show a notable change between baseline and the follow-up (Baseline: 6.8± 3.7; Follow-up: 7.1± 4.1, BF_10_: 0.255), indicating the persistent nature of this core feature of schizophrenia.

To study the longitudinal trajectory of word usage during descriptive discourse, we performed a Bayesian paired t-test from baseline to 6 months follow up in both groups. As shown in Table 3, the null model was more likely than the difference-between-measures model for the HC group across all the linguistic variables, indicating relative stability of semantic co-occurrence and the number of produced words among healthy subjects, when the same pictures were described twice in a period of ∼6 months. In the SZ group, the most notable evidence for the difference between measures was noted for ASW-F. The evidence for a longitudinal increase in ASW-F over 6 months was nearly 6.3 times compared to the null model of no change in patients (Figure 1). We did not see the same level of evidence for linear change in ASW-10 or number of words. For further correlational analysis with cognitive and symptom factors, we selected ASW-F as the linguistic measure of interest.

**Table 3.**
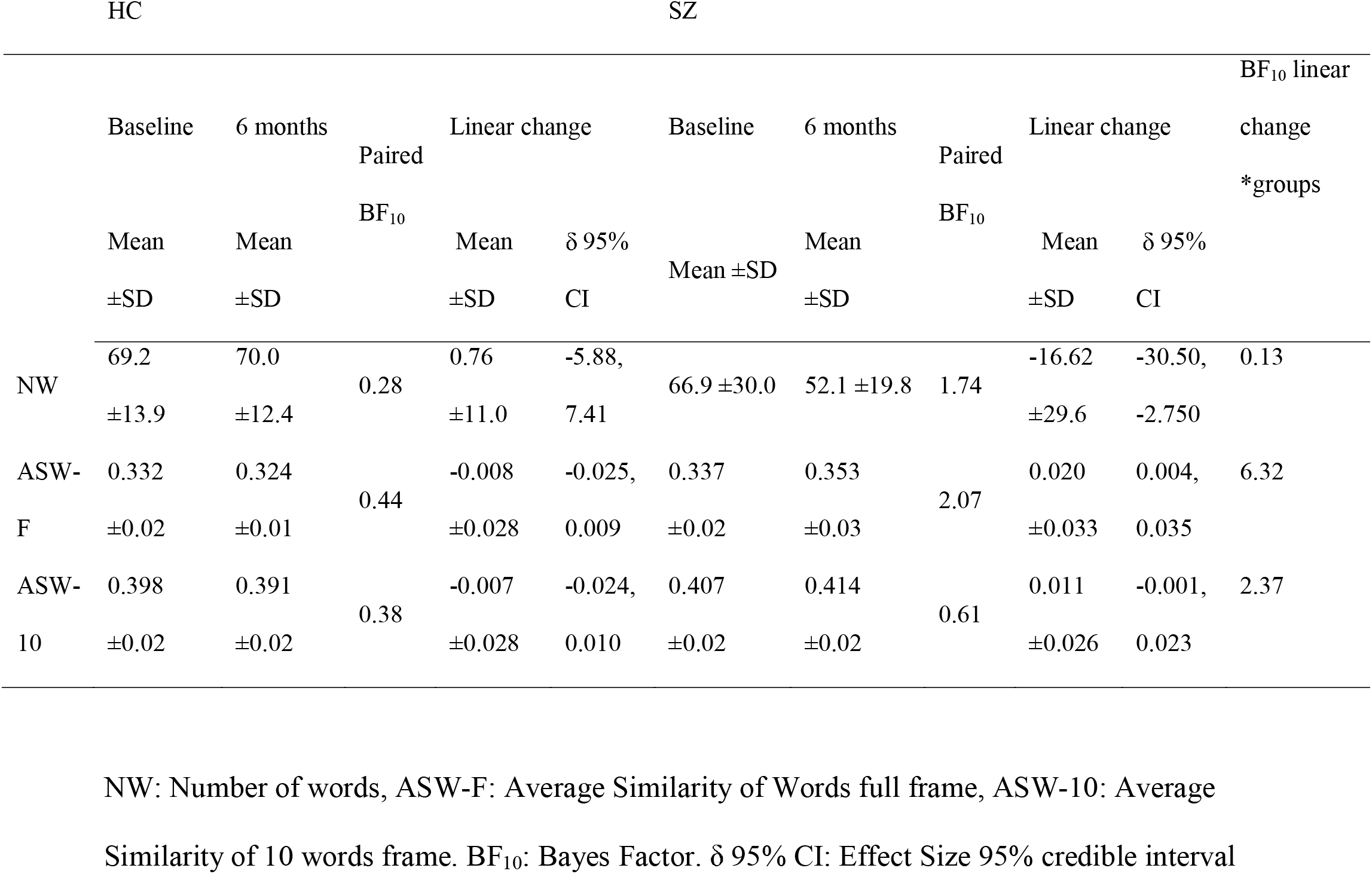
Summary of baseline and follow up 6 months comparison.

### Symptoms, functioning, and word similarity

Among FES subjects, ASW-F at the time of illness onset was higher in the presence of more severe positive symptoms (PANSS-8 positive r: 0.39, BF_10_: 9.24) and reduced functioning (SOFAS scores r: -0.41, BF _10_: 128) but this relationship was not seen with PANSS-8 negative (r: 0.08, BF_10_: 0.18) scores, TLI impoverishment (r: 0.21, BF_10_: 0.49), disorganization (r: 0.14, BF_10_: 0.28) or dysregulation (r: -0.06 BF_10_: 0.20) scores (Figure 2). Among FES subjects that were followed-up, there was moderate evidence for increasing ASW-F in patients with increasing PANSS-8 negative (r: 0.592, BF_10_: 18.7) but not with change in PANSS-8 positive (r: -0.125 BF_10_: 0.435), or SOFAS scores (r: -0.04 BF_10_: 0.322).

**Figure 2.**
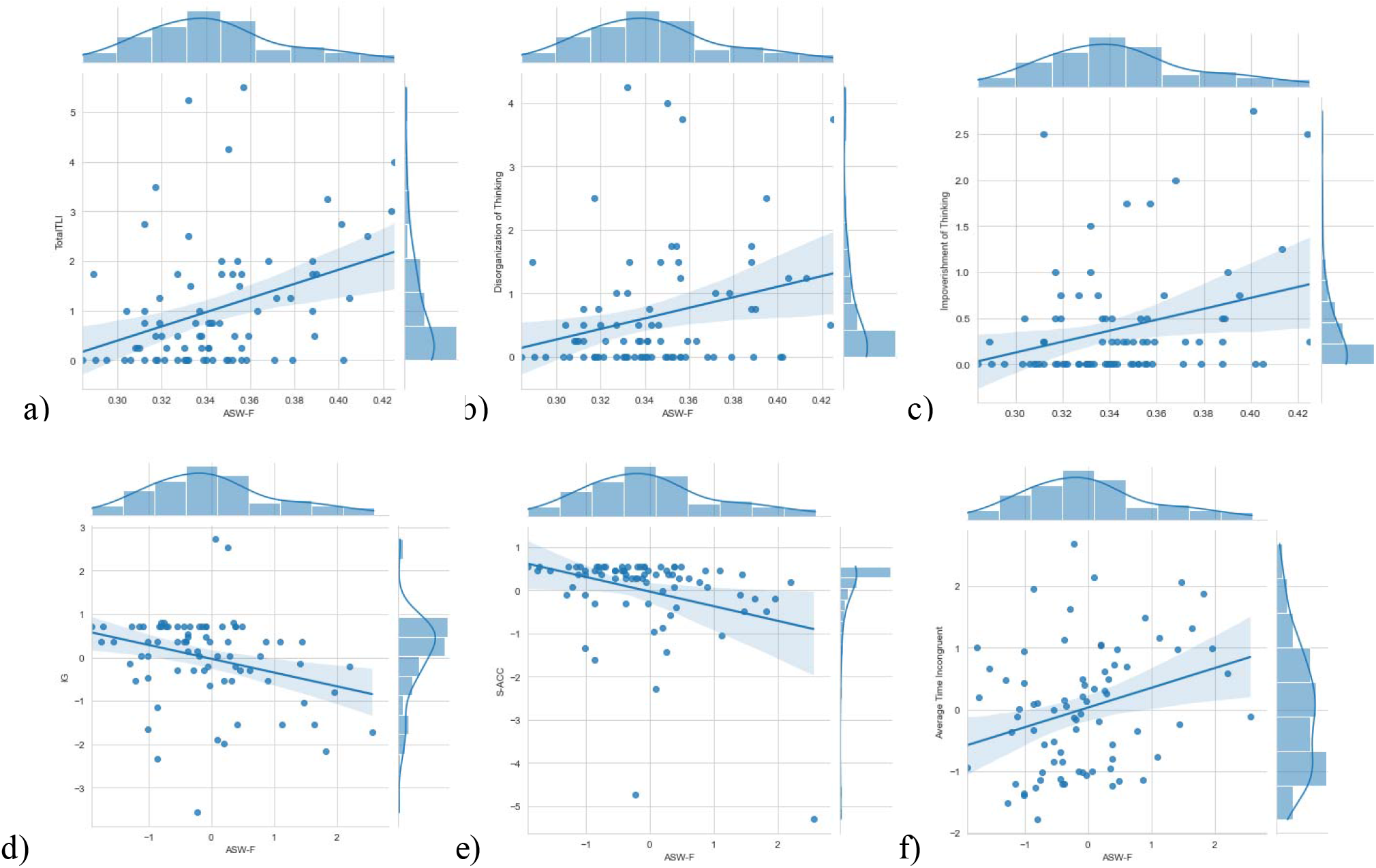
Correlation between ASW-F and TLI symptoms. ASW-F: Average similarity of Words full frame with a) Total TLI (Thought Language Index), b) Disorganization of thinking and c) Impoverishment of thinking, d) IG: Interference score, e) S-ACC: Stroop accuracy and f) Response time incongruent condition.

### Cognition and word similarity

When all subjects (patients and controls) at the baseline were considered together, ASW-F was higher in subjects with reduced Stroop accuracy (r: -0.31, BF_10_: 13.3) The within-group effects were weaker, but in the same direction (FES only: r: -0.22, BF_10_: 1.01; HC only: r: -0.29, BF_10_: 1.61). Higher ASW-F cores also related to lower Interference score (of Golden: IG) (r: -0.29, BF_10_ of 8.24, ; FES only: r: -0.20, BF_10_: 0.81; HC only: r: -0.25, BF_10_: 1.13) and prolonged reaction time for the incongruent condition (r: 0.29, BF_10_: 8.6; FES only: r: 0.28, BF_10_: 1.97; HC only: r: 0.06, BF_10_: 0.29). This indicates that semantic co-occurrence in discourse production was higher in the presence of a cognitive control deficit indexed by reduced inhibitory control (poor accuracy) and information processing speed. A more specific index of serial processing speed, DSST, was also lower in the presence of increased ASW-F in the entire sample (r: -0.48, BF_10_: 304). This association was largely driven by the FES group (r: -0.41, BF_10_: 7.99), not the HC (r: - 0.03, BF_10_: 0.21) (see more details in the supplementary materials).

### Effect of antipsychotics exposure

To investigate possible effects of antipsychotics, we related both the Daily dose (average Daily Defined Dose) and Total Dose (total exposure calculated based on Daily Dose and number of days of exposure) to NW and ASW-F at both time points. As shown in Table 4, the difference between the baseline and follow up measures on NW and ASW-F were not correlated with Daily Dose or Total Dose.

**Table 4.** Relationship between 6-months change in linguistic variables and medication dose.

|  | Pearson's r | BF<sub>00</sub> | Lower 95% CI | Upper 95% CI |
| --- | --- | --- | --- | --- |
| Daily Dose - NW | 0.105 | 0.303 | -0.330 | 0.491 |
| Daily Dose- ASW-F | -0.161 | 0.343 | -0.530 | 0.283 |
| Total Dose - NW | 0.083 | 0.293 | -0.348 | 0.475 |
| Total Dose- ASW-F | -0.225 | 0.424 | -0.574 | 0.227 |

Daily dose = average Daily Defined Dose, Total Dose: total exposure calculated based on Daily Dose and number of days of exposure. NW - Number of words. ASW-F: Average Similarity of Words full frame.

### Effect of social factors on word similarity

To investigate possible effects of immigrant status and the use of language other than English at home^48^, we removed 20% subjects that satisfied this criterion, and analysed the difference in ASW-F at baseline. We continued to see evidence in favour of increased ASW-F among patients with FES (ASW-F BF_10_ = 6.46). Similarly, when patients were stratified according to education status (<12/>12 years) and by parental socio-economic status (higher than median vs. lower than median) were compared with each other, there was no difference in ASW-F, ASW-10 or NW (Educational background: ASW-F BF_10_ = 0.594, ASW-10 BF_10_ = 0.581, NW BF_10_ = 0.173; Socio-economic status: ASW-F BF_10_ = 0.194 ASW-10 BF_10_ = 0.179, NW BF_10_ = 0.148). These results indicate that word similarity is affected by diagnosis of schizophrenia per se, rather than the social factors that are often associated with diagnosis.

## 3. Discussion

Using a computational semantics approach, we examined word similarity during a controlled descriptive discourse task in untreated first episode schizophrenia at baseline and after 6 months of treatment. We report three major findings. First, when faced with the task of providing a description of an unfamiliar concrete referent (a picture), patients with schizophrenia choose words with higher probability of semantic co-occurrence. The likelihood of this phenomenon is more pronounced when psychotic symptoms are severe and functional deficits are profound. Interestingly, this objectively verifiable linguistic feature of higher similarity is seen irrespective of the degree of clinically detectable thought disorder. Second, higher word similarity during the discourse related to lower cognitive control, as indexed by Stroop task, and reduced processing speed, indicating a role for domain general processes in aberrant word choices in schizophrenia. Third, despite symptomatic improvement with treatment (i.e. reduction of positive symptoms), the higher similarity of words used for descriptive purposes worsened with time among patients. This suggests that the restricted sampling from the semantic space is a specific deficit, associated with but not fully explained by the acuity of symptoms and functional deficits, that does not respond to dopaminergic early intervention but follows the trajectory of negative symptoms. Taken together, these three findings imply that language processing anomalies may play a key role in the longitudinal trajectory of psychosis; understanding the mechanisms behind these disruptions may provide a window to reverse a key factor contributing to persistent deficits among patients.

Semantic impairment in people with schizophrenia is widely reported ^49^, however, this evidence relies mostly on comprehension based experimental paradigms ^50–52^ or experiments where the semantic retrieval demand, or route in the semantic space, is set by the researchers (stimulus with prime and target) and not chosen by the participants. Studies of the latter type generally involve category fluency tests, wherein patients have either no reduction in overall word similarity or choose adjacent words that are less similar^26,41^. In contrast to verbal fluency tasks, in a discursive task there is a necessity to ‘forage’ widely to accomplish the goal of description. Such wide foraging appears to be diminished in schizophrenia^53^. We also note that such a narrowing of semantic sampling space relates to higher Stroop interference effect; thus, a failure of the prefrontal executive control, either in a general- or domain-specific manner ^54^, may influence the word choices. The lack of control in the selection of the lexical itemsClick or tap here to enter text. may lead to a restricted repertoire wherein a word and its activated associates ^56,57^ dominate the unfolding discourse.

Our study has several strengths as well as some limitations. To our knowledge, this is the first longitudinal report on the nature of word choices made during a discourse in psychosis. Although the evolution of language or semantics in schizophrenia is still not fully understood, meta-analytical evidence indicates no temporal change when category fluency is tested - indicating its fixed, endophenotype-like stability over time ^58^. In contrast, we report the discourse-specific word choice deteriorates over time in early stages of schizophrenia. Secondly, we estimated antipsychotic exposure meticulously over the follow-up period. The discourse-related word similarity did not change in proportion to antipsychotic dose exposure, in contrast with the reported influence of antipsychotic dose on other NLP measures such as syntactic complexity and percentage of time speaking^23^. We were limited in terms of the number of healthy controls for whom we had follow-up assessment of word similarity; nevertheless, this did not diminish our ability to demonstrate group differences in the longitudinal change scores based on within-subject variance. Secondly, our descriptive discourse was constrained by time; we do not know if the choice of words would have been less similar if the discourse was unconstrained and spontaneous. This needs to be examined in future studies with speech elicited in different contexts.

In conclusion, we demonstrate that descriptive discourse in first episode of schizophrenia group is characterised by an aberrantly high semantic co-occurrence that relates to functional deficits and progressively worsens in early stages. Given its relationship with poor functioning, our ability to measure it objectively and repeatedly in a non-invasive manner, we propose this measure to be a suitable treatment target that indexes the core, hitherto unclear, progressive pathology of schizophrenia.

## 4. Methods

### Participants

Eighty-two English-speaking participants were recruited, including 46 with First Episode of Schizophrenia (FES) and 36 healthy controls (HC). FES participants were enrolled through the Prevention and Early Intervention for Psychosis Program of London Health Sciences Centre (Canada) and were diagnosed with Schizophrenia according to the DSM-5 criteria, using a consensus procedure that confirmed diagnosis 6 months after the first presentation^59^. Severity of symptoms was confirmed with the Positive and Negative Syndrome Scale-8 items version (PANSS) ^60^. The FES participants were drug-naïve for antipsychotics at the time of assessment (total antipsychotic use equal or less than 14 days).

The HC group recruitment criteria included no personal or family mental illness or neurological diseases. The groups were matched in age, sex and level of completed formal education. Participants were assessed with the Social and Occupational Functioning Assessment Scale (SOFAS) which is a rating scale of functioning level with emphasis on the social and occupational aspects. The SOFAS scores the level of functioning in interpersonal, occupational and self-care roles, without overlapping with symptom measurements ^61^. The FES group was assessed with the Calgary Depression Scale (CDS) ^62^ a Clinical Global Impression Scale Severity of Illness (CGI-S) ^63^. All participants provided written informed consent before assessment and ethics approval was granted by the Human Research Ethics Board at Western University, London, Ontario.

Thirty participants, 20 with schizophrenia (SZ) and 13 HC, were followed up approximately 6 months from the first assessment (x□ = 214.9 ± 44.9 days). The medication exposure of the SZ group was calculated according to the Daily Defined Dose (DDD) methodology ^64^. To calculate total exposure, we considered the type of medication, the dose prescribed, the number of days of effective exposure based on treatment compliance over the follow-up time measured using an established instrument^65^ for adherence that correlates well with pill counts^66^. As reported in our prior study^67^, nearly 50% of patients went on long-acting injection by the 1^st^ month of treatment, further ensuring treatment compliance.

### Assessment

Participants were cognitively assessed using the digit symbol substitution task oral and written version (DSSTo and DSSTw), Semantic verbal fluency and Stroop test. The DSST oral and written version was scored counting the number of correct symbols within the allowed time, total DSST was calculated with oral and written version average. For the Semantic verbal fluency task, participants were instructed to say the maximum number of animals in one minute. We analysed the number of words as the mean similarity between words with the Covington Vector semantic tool. In the Stroop test the performance was measured by number of correct answers (S-ACC), the response time in incongruent condition and the Interference score (IG). The IG was calculated with Golden method ^68^, in which the predicted color-word (pCW) is the product of the word(W) and color (C) scores with the following formula:

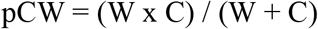

Then, the interference score (IG) was computed subtracting the pCW from the incongruous condition (CW) as follows ^69^:

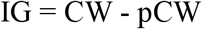

The discourse task was the description of 3 images and the scoring was done using the Thought Language Index (TLI). The TLI is a reliable instrument used to assess formal thought disorders under standardized conditions ^3^. The participants are asked to describe Thematic Apperception Test^70^ images and are given one minute for each image. The interviewer prompts the participants if they stop speaking before the time is over. The interview is recorded and later transcribed by research assistants. The transcriptions are then analysed with the Covington Vector semantic tool^71^.

### Semantic Analysis

The Covington Vector semantic tool (CoVec) is a natural language processing tool based on data from Global Vectors for Word Representation (GloVe) Project, with 840 billion words in English on a 300-element vectors^72^. GloVe measures the likelihood of co-occurrence of words through vector cosine similarity based on overall statistics of how often the word appear given the context (P(w|c)). The GloVe project is count-based model with a large matrix of (words*context) co-occurrence information that is normalized by log-smoothing the matrix. Covec reports the average of similarity, that is, whether successive words are commonly used in the same context (or together), with an n-word frame segment, using all the positions of the frame. Before processing the text, CoVec removes punctuation, marks ‘stop words’ (eg. “a”, “the”, “is”, “at”, among others), and finally, ignores words which are not found in the GloVe dataset (displays a warning of all the missing words). The metrics used include the Number of words (NW), Average Similarity of Words (ASW), summarized as Coherence in the tool, ASW in the full frame of the text (ASW-F) and ASW in 10 words moving window (ASW-10).

### Data analysis

Clinical and demographic data were analysed using descriptive and Bayesian statistics. We first compared group performance with a Bayesian t test on the NW and ASW variables. In order to compare the progression of language features, we conducted a Bayesian paired t test between baseline and 6 months follow up measures, then, we estimate the linear change between measures and compare between groups. We conducted a Bayesian Pearson correlation to explore the effect of antipsychotics on our language variables. To address the interaction with cognitive and symptoms variables, a Bayesian correlation were made between semantic co-occurrence and Stroop, DSST, TLI and PANSS. The variables were correlated considering the linear change between baseline and follow up and were standardized by dividing the linear change with the baseline. Finally, we test the effect of the use of language other than English, educational background and socio-economic status of the parents with Bayesian t test for two groups stratification and Bayesian ANOVA for three groups stratification. The prior distribution for the parameter was set by default. All the statistical analysis used JASP version 0.14.0.1^73^ and the figures were made on Python in Jupyter Notebook 6.1.5^74^.

## Data Availability

Data is available upon request

## Acknowledgements

We appreciate all the participants and their families for the time and effort to contribute to this study. We are grateful to Peter Jeon (Robarts Research Institute) for Stroop task data acquisition. We thank Michael Covington (Covington Innovations) for providing us his CoVec NLP tool. We thank all research team members of the NIMI lab and all the staff members of the PEPP London team, particularly Drs. Kara Dempster (currently at Dalhousie University), Julie Richard, Priya Subramanian and Hooman Ganjavi for their assistance in patient recruitment and supporting clinical care.

## Author Contributions

MFA and LP conceptualized the project, SF and MM collected the data, MF analysed the data, MF and LP wrote the manuscript; RL, AS and all authors critically reviewed and approved the final version of the manuscript.

