## supplemental figure 2 for "Progressive changes in descriptive discourse in First Episode of Schizophrenia: A longitudinal computational semantics study"

Supplementary materials

Correlation between ASW-F and Stroop ACC

FES	HC	All

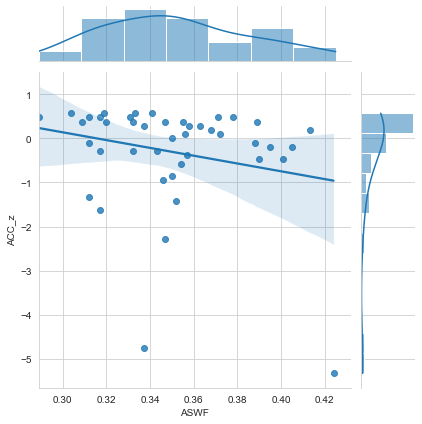

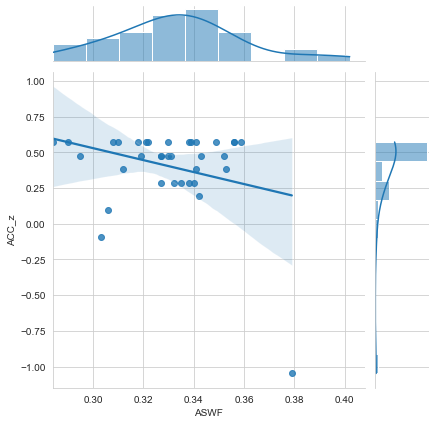

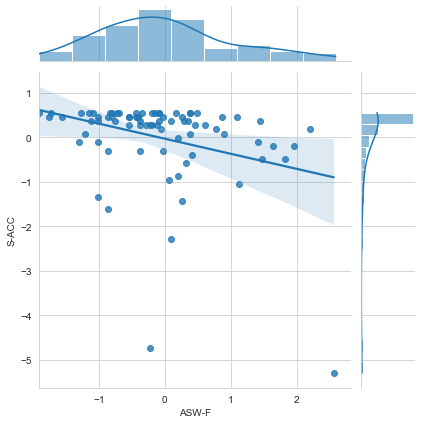

Correlation between ASW-F and Stroop IG

FES	HC	ALL

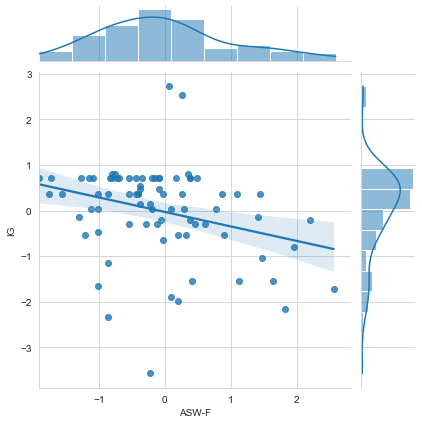

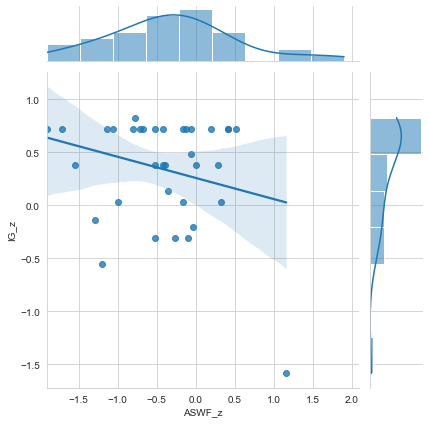

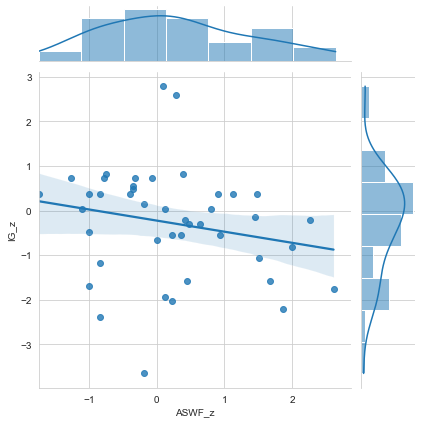

Correlation between ASW-F and Stroop total time incongruent condition

FES	HC	All

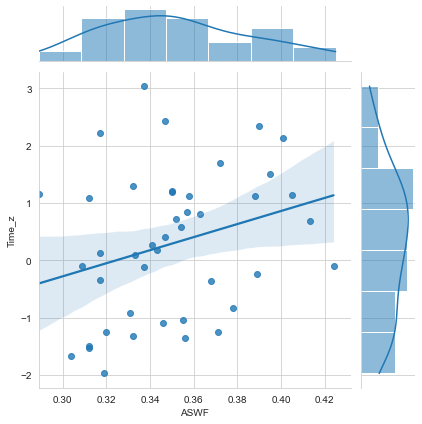

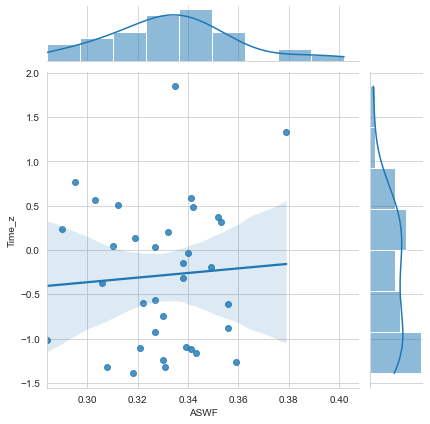

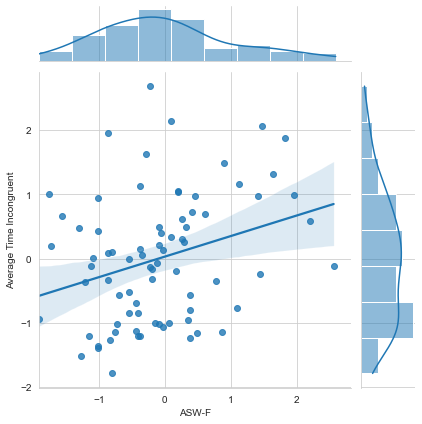

Correlation between ASW-F and DSST

FES	HC	All

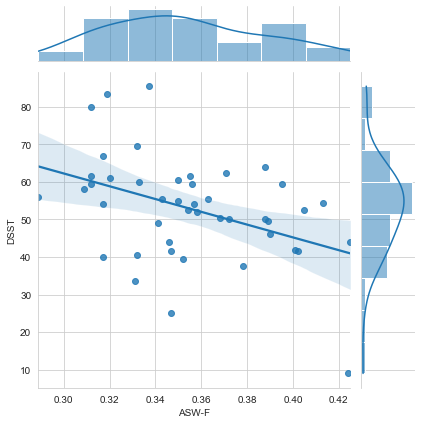

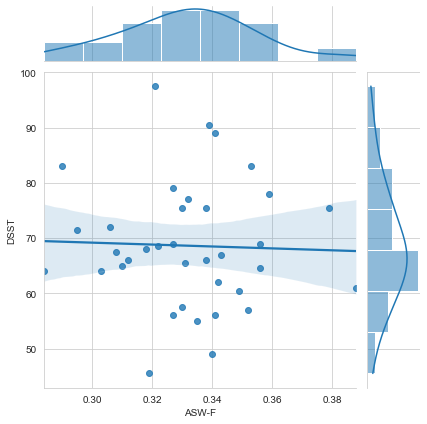

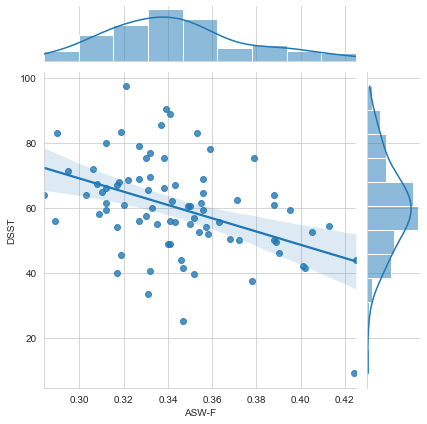

Correlation between ASW-F and DSSTw

FES	HC	All

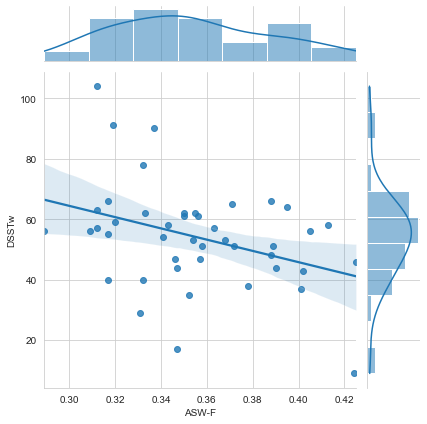

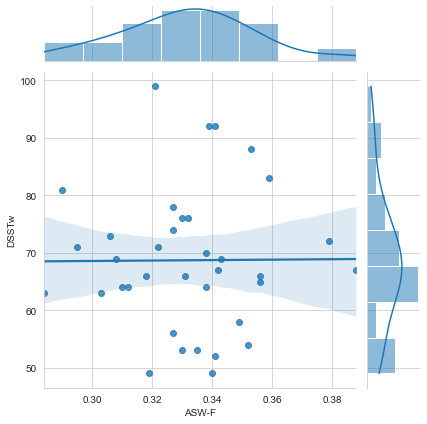

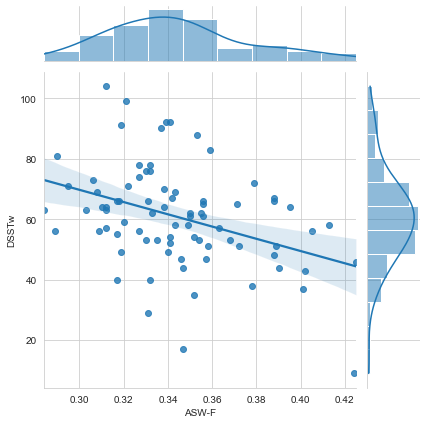

Correlation between ASW-F and DSSTo

FES	HC	All

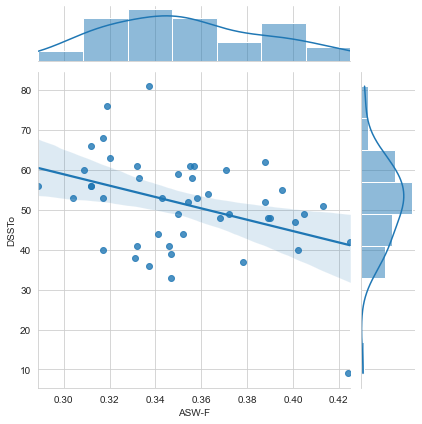

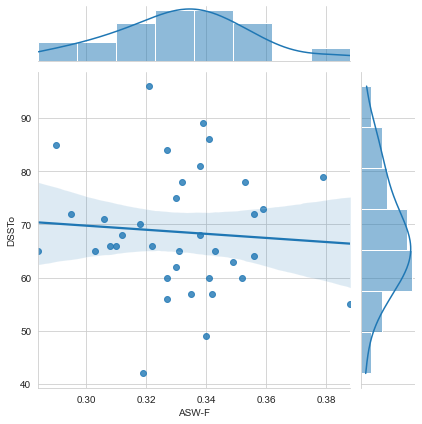

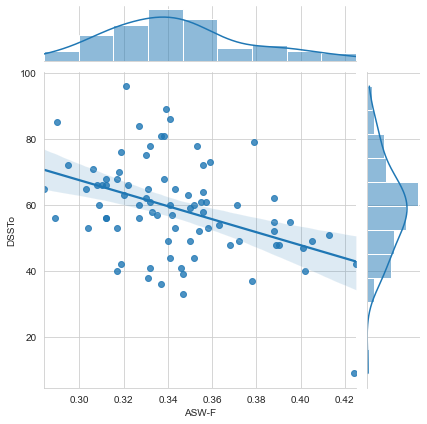
